# Machine Learning for Paediatric Related Decision Support in Emergency Care – A UK and Ireland Network Survey Study

**DOI:** 10.1101/2025.06.29.25330501

**Authors:** Fiona Leonard, Mark D Lyttle, Dympna O’Sullivan, John Gilligan, Damian Roland, Michael Barrett, PERUKI

## Abstract

This study explores clinician understanding and perception at site lead level towards machine learning (ML) decision support tools for paediatric related emergency care across the UK and Ireland, essential in guiding safe and effective frontline implementation. A cross-sectional online survey was distributed via Paediatric Emergency Research United Kingdom and Ireland (PERUKI) to the lead for digital systems or PERUKI site lead, with one response sought per site. Survey development was in REDCap, and descriptive analysis (counts, percentages) was performed. The response rate was 86.7% (65/75), mostly from England (83.1%). While 80.0% understood ‘Artificial Intelligence’, fewer understood advanced concepts such as ‘Deep Learning’ (32.3%). Most clinicians believed ML will support decision making (83.1%), would be willing to use (87.7%), and the future of decision making is a combination of human and ML (83.1%). Barriers included concerns about bias (61.5%), ML accuracy (56.9%), and inadequate information technology infrastructure (67.7%). Digital leads were more concerned about ML accuracy than non-digital (68.2% vs. 51.2%). Among potential applications, antimicrobial stewardship ranked highest (90.8%), and diagnosis of mental health conditions lowest (24.6%). Strong interest in ML tools for decision support in paediatric emergency care was evident, though concerns about bias, accuracy, and infrastructure must be addressed. Ongoing co-design with clinicians is critical in ensuring these tools are trusted, useful and suited to paediatric emergency care. Targeted education, digital leadership, and strategic investment in infrastructure and governance are essential for the successful adoption and integration of ML in clinical workflows.

**Author Summary:** Within emergency care we are seeing a rapid growth in the research, development and frontline use of machine learning based tools for decision support, yet very little is known about the intended users understanding, opinions, experience of this technology and supporting structures. With any new technology a greater understanding leads to better adoption and the clinicians who will use these tools should be directly involved in their design, implementation and evaluation to ensure that these tools are clinically relevant and usable in practice. Through our survey of clinical site leads (key drivers, influencing the adoption of ever advancing technology), the findings provide critical insight into what clinicians are most concerned about, their perceptions, understanding, what applications they view as most clinically relevant and their willingness to be involved in future research and development of these tools. The results also revealed many important themes such as infrastructure readiness, trust, explainability, clinical integration, targeted education and human-artificial intelligence collaboration. These findings will contribute to shaping the future of research, development, education, governance and policy within this rapidly growing area.

## Introduction

Despite increasing research on machine learning (ML) for clinical decision support, its understanding, adoption, use, and perception among emergency department (ED) clinicians remain underexplored. The rapid growth of ML, fuelled by electronic health records (EHRs) [1], highlights the need for further study in ED settings. The potential application of these tools are numerous, supporting decision making from when the patient arrives in the ED, during that visit and through to discharge [2] (e.g. quickly predicting triage, risk stratifying the most unwell patients [3], artificial intelligence (AI) assisted radiology diagnosis [4], and predicting discharge outcomes for bed planning [5,6]). However, obstacles to adopting into real-world practice are abundant, with concerns including explainability, data bias, socio-technical, ethical, workforce, liability, patient safety, and the ability to integrate into clinical workflows [7–9]. The role of the institutional and departmental digital leads is integral to the adoption of these technologies.

Digital leaders take a more proactive role in advancing their organisations, supporting the adoption of modern and ever evolving digital healthcare technologies [10]. Digital leadership can positively transform attitudes and anxiety towards AI [11], therefore this role is pivotal in fostering greater acceptance, confidence, and engagement in the integration and use of ML decision support tools.

This study sought to assess the knowledge, perceptions, and obstacles to the adoption of ML based decision support tools among digital (if the position existed) and non-digital clinical leaders in paediatric emergency care across the United Kingdom and Ireland. Through surveying both digital and non-digital leaders, the study aimed to capture perspectives on ML integration, key challenges, and the crucial role of digital leadership which is instrumental in driving AI and ML adoption.

### Key concepts

- **Artificial Intelligence:** The ability of computers or machines to mimic intelligent behaviour, generating results that were once believed to require human intelligence [12].
- **Machine Learning:** A sub field of Artificial Intelligence. The ability of computers to learn and adapt on their own by analysing patterns in data using algorithms and statistical models, without needing explicit instructions [13].
- **Knowledge Based Systems:** These systems follow explicit instructions, typically structured as IF-THEN rules, where data is retrieved and assessed to determine the appropriate output or action [14].
- **Non-knowledge Based Systems:** These systems leverage Artificial Intelligence, Machine Learning or statistical models trained on datasets enabling computers to learn from past experiences and find patterns in data to provide outcomes, rather than being explicitly programmed [14].

## Materials and methods

A cross-sectional online voluntary and anonymised survey study was conducted. The survey questions were primarily informed by previous research [15–20] and refined through input, discussion and consensus among the study investigators (survey sections in Table 1), several of whom are extensively experienced in survey design. Survey usability and technical testing was undertaken by the study investigators and an independent clinician reviewer, with revisions made based on feedback received. The survey questions are detailed in S1 File.

**Table 1.** Survey sections, with number of questions and answer types.

| Survey Section | Number of questions | Types |
| --- | --- | --- |
| Study participant information | 6 | Multiple choice, single answer |
| Site and participant demographics | 5-9 | Multiple choice, drop-down list<br>Text<br>Multiple choice, Single answer<br>Multiple choice, multiple answer |
| Confidence in understanding key artificial intelligence concepts | 6-11 | Likert (5 values)<br>Text<br>Multiple choice, single answer |
| Perceptions, concerns, and the future of machine learning for decision support | 16 | Likert (5 values) |
| Current information technology set up and current experience with clinical decision support system tools | 9-18 | Multiple choice, Single answer<br>Text<br>Multiple choice, multiple answer |
| Reasons machine learning decision support tools are not used in their clinical setting | 0-10 | Likert (5 values)<br>Text |
| Training in machine learning | 3-4 | Multiple choice, Single answer<br>Text<br>Multiple choice, multiple answer |
| Areas of application for machine learning in emergency medicine | 24-25 | Likert (5 values)<br>Text |
| Data repository with anonymised patient data for research purposes | 5 | Likert (5 values)<br>Multiple choice, Single answer |

The online survey was developed in Research Electronic Data Capture (REDCap) [21,22], a secure online software platform designed to facilitate data collection for research. Branching logic was used on some questions which varied the number of questions asked depending on the responses provided. For sections with many questions, random order was utilised to reduce study fatigue and enhance the viability of the responses. All except two open questions (branching logic dependent) were mandatory. Anonymised data was stored securely on a server in Technological University Dublin (TU Dublin), accessible to the study investigators only.

An educational video (3.5 minutes) was embedded to explain key decision support types and AI concepts (https://vimeo.com/892770833/180bc711e1<u>?</u>). Participants were initially asked five questions to assess their understanding of key AI concepts before watching the video. After viewing the video, they were asked if they were satisfied with their initial understanding. Among those who responded negatively, the five questions were repeated.

The study was delivered across Paediatric Emergency Research in the United Kingdom and Ireland (PERUKI). Sites comprise a range of types and locations, including stand-alone paediatric and mixed adult/paediatric ED, in urban and rural settings. The survey link was distributed to one person at each of the 75 PERUKI sites; where a digital lead existed, they were the recipient, and where there was no digital lead, the response was provided by the PERUKI site lead. The duration of the survey was four weeks beginning on 14^th^ June 2024 and ending on 12^th^ July 2024. Three reminders were sent, two weeks and three weeks after opening and 48 hours before the close of the survey.

Data exported from REDCap was prepared for analysis by checking and adjusting formats, removing incomplete surveys, adding summary columns, and verifying data integrity in both REDCap and Microsoft Excel. Descriptive analysis, including counts, percentages, and chart visualisation, was conducted in Microsoft Excel. Percentages shown in the results for Likert scales, summed ‘Agree’ and ‘Strongly Agree’ together, ‘Disagree’ and ‘Strongly Disagree’ and likewise summing ‘Likely’ and ‘Extremely Likely’ where applicable. The study adhered to The Checklist for Reporting Results of Internet E-Surveys (CHERRIES) guidelines [23], with the completed checklist provided in S2 File.

### Ethical approval and participant consent

Ethical approval for this survey was received from both Children’s Health Ireland (REC-281-23) and TU Dublin (LEWS-023-83) Research Ethic Committees. Prior to gaining access to the survey, participants were required to provide electronic (written) informed consent by affirming six mandatory statements regarding their understanding of the study, the terms and their agreement to participate (S1 File).

## Results

Only completed responses were included in the results. The survey response rate was 86.7% (65/75 sites) after thirteen incomplete or duplicate responses were excluded. Most sites were in England (83.1%, 54/65) and treated both paediatric and adult patients (40/65, 61.5%). The digital lead position existed in 60.0% (39/65) of departments, and 83.1% (54/65) utilised healthcare records to drive improvements. Patient records were predominantly recorded electronically (Table 2).

**Table 2.** Site Characteristics.

| **Characteristic** | **Value (n=65)**<br>**n (%)** |
| --- | --- |
| **Country/Constituent Country** |  |
| England | 54 (83.1) |
| Republic of Ireland | 6 (9.2) |
| Scotland | 3 (4.6) |
| Wales | 1 (1.5) |
| Northern Ireland | 1 (1.5) |
| **Site Type** |  |
| Paediatric and Adult | 40 (61.5) |
| Paediatrics | 25 (38.5) |
| **Does a digital lead position exist in your department?** |  |
| Yes | 39 (60.0) |
| No | 26 (40.0) |
| **Does your department utilise data from healthcare records to drive improvements?** |  |
| Yes | 54 (83.1) |
| No | 11 (16.9) |
| **Emergency department patient records are recorded on** |  |
| Fully electronic | 33 (50.8) |
| Paper and electronic system | 29 (44.6) |
| Paper based | 3 (4.6) |
| **Type of Information System** |  |
| Emergency Department module within a hospital wide EHR System | 28 (43.1) |
| Integrated Emergency Department Information System | 20 (30.8) |
| Standalone Emergency Department Information System | 10 (15.4) |
| Hybrid System | 7 (10.8) |

The role of digital lead was held by 33.8% (22/65) respondents; however, 17/65 respondents completed the survey in place of the digital lead. The reasons provided included the absence of a designated paediatric emergency medicine digital lead, the role being covered by adult ED lead, paediatric or non-clinical leads. In some sites this role was temporarily vacant or shared among many team members.

Respondents were predominantly Consultants, representing 95.5% (21/22) of digital leads and 97.7% (42/43) of non-digital leads. The majority had at least six years of experience in emergency medicine (digital leads: 77.3% (17/22), non-digital leads: 88.4% (38/43)). Most respondents (40.0%, 26/65) had paediatrics with PEM subspecialty training (Table 3).

**Table 3.** Respondent Characteristics.

| **Characteristic** | **All Positions**<br>**(n=65)**<br>**n (%)** | **Digital**<br>**(n=22)**<br>**n (%)** | **Non-digital**<br>**(n=43)**<br>**n (%)** |
| --- | --- | --- | --- |
| **Years Experience** |  |  |  |
| 0-2 | 1 (1.5) |  | 1 (2.3) |
| 3-5 | 9 (13.8) | 5 (22.7) | 4 (9.3) |
| 6-10 | 17 (26.2) | 5 (22.7) | 12 (27.9) |
| 10+ | 38 (58.5) | 12 (54.5) | 26 (60.5) |
| **Role** |  |  |  |
| Consultant | 63 (96.9) | 21 (95.5) | 42 (97.7) |
| Consultant Nurse | 1 (1.5) |  | 1 (2.3) |
| Paediatric Registrar | 1 (1.5) | 1 (4.5) |  |
| **Clinician Type** |  |  |  |
| Paediatrics with PEM subspecialty training | 26 (40.0) | 9 (40.9) | 17 (39.5) |
| Emergency Medicine | 17 (26.2) | 7 (31.8) | 10 (23.3) |
| Emergency Medicine with PEM subspecialty training | 14 (21.5) | 3 (13.6) | 11 (25.6) |
| Paediatrics | 6 (9.2) | 2 (9.1) | 4 (9.3) |
| Unknown | 2 (3.1) | 1 (4.5) | 1 (2.3) |

After the video explaining AI concepts, 41.5% (27/65) revised their answers (digital leads: 31.8% (7/22), non-digital leads: 46.5% (20/43)). Digital leads understood all key concepts better pre-video, but post-video, non-digital leads outperformed them in 3/5 concepts. The highest level of understanding pre-video was for the concept of ‘Artificial Intelligence’ at 80.0% (52/65), which increased post-video to 89.2% (58/65). Confidence in deep learning rose for both groups, from 32.3% (21/65) to 56.9% (37/65), with non-digital leads seeing the biggest gain (+27.9% vs. +18.2% for digital leads). Similarly, natural language processing understanding improved from 46.2% (30/65) to 69.2% (45/65), with non-digital leads again showing greater understanding (+25.6% vs. +18.2%) (Fig 1).

**Fig 1.**
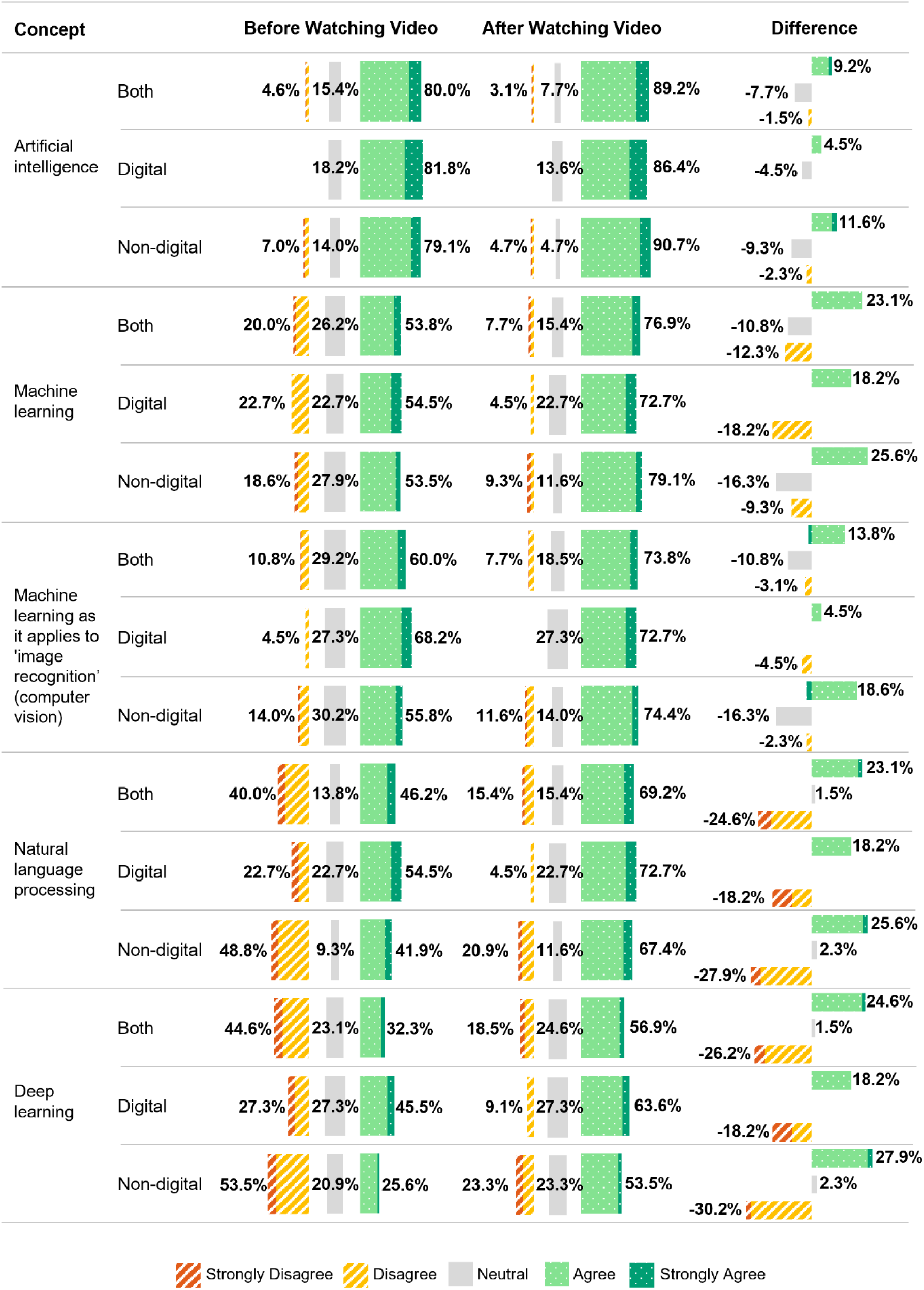
Confidence in Understanding Key Artificial Intelligence Concepts by digital and non-digital lead group, pre and post watching educational video.

Knowledge-based (rule based) decision support tools are more widely used than non-knowledge-based (ML) tools (38.5% (25/65) compared to 10.8% (7/65)). Among digital leads, 45.5% (10/22) used knowledge-based tools, compared to 34.9% (15/43) of non-digital leads. Only 10.8% (7/65) reported using non-knowledge-based tools (Table 4).

**Table 4.** Use of knowledge and non-knowledge based tools.

|  | Knowledge based<br>(Rule based) |  |  | Non-knowledge based<br>(Machine Learning) |  |  |
| --- | --- | --- | --- | --- | --- | --- |
|  | Digital<br>(n=22)<br>n (%) | Non-digital<br>(n=43)<br>n (%) | Both<br>(n=65)<br>n (%) | Digital<br>(n=22)<br>n (%) | Non-digital<br>(n=43)<br>n (%) | Both<br>(n=65)<br>n (%) |
| Yes | 10 (45.5) | 15 (34.9) | 25 (38.5) | 4 (18.2) | 3 (7.0) | 7 (10.8) |
| No | 10 (45.5) | 22 (51.2) | 32 (49.2) | 18 (81.8) | 40 (93.0) | 58 (89.2) |
| Unsure | 2 (9.1) | 6 (14.0) | 8 (12.3) |  |  |  |

Most respondents expressed willingness to use ML decision support tools (87.7%, 57/65) and 83.1% (54/65) believed these tools would support decision making. The majority agreeing these tools should be developed in response to clinical need (87.7%, 57/65) and should consider the broader socio-technical requirements (95.4%, 62/65). There was strong agreement that trust in these tools is underpinned by explainability (86.2%, 56/65). However, 67.7% (44/65) had significant concerns around infrastructure readiness and skilled resources, 56.9% (37/65) were concerned about ML accuracy and 53.8% (35/65) believed these tools may impact the ability for clinicians to accurately assess patients. Opinions on clinical risks and biases were divided, with 40.9% (9/22) of digital leads and 30.2% (13/43) of non-digital leads expressing concerns about increased risks and 59.1% (13/22) of digital leads and 62.8% (27/43) of non-digital leads raising concerns about potential biases. Overall, clinicians support integrating ML into emergency medicine, agreeing the future will be a combined human-ML approach (83.1%, 54/65). Non-digital leads (88.4%, 38/43) were more optimistic than digital leads (72.7%, 16/22), and a large proportion advocated its inclusion in medical training curricula (digital leads: 77.3% (17/22), non-digital leads: 67.4% (29/43)). Most (81.5%, 53/65) expressed interest in contributing to future research and development in ML decision support (Fig 2).

**Fig 2.**
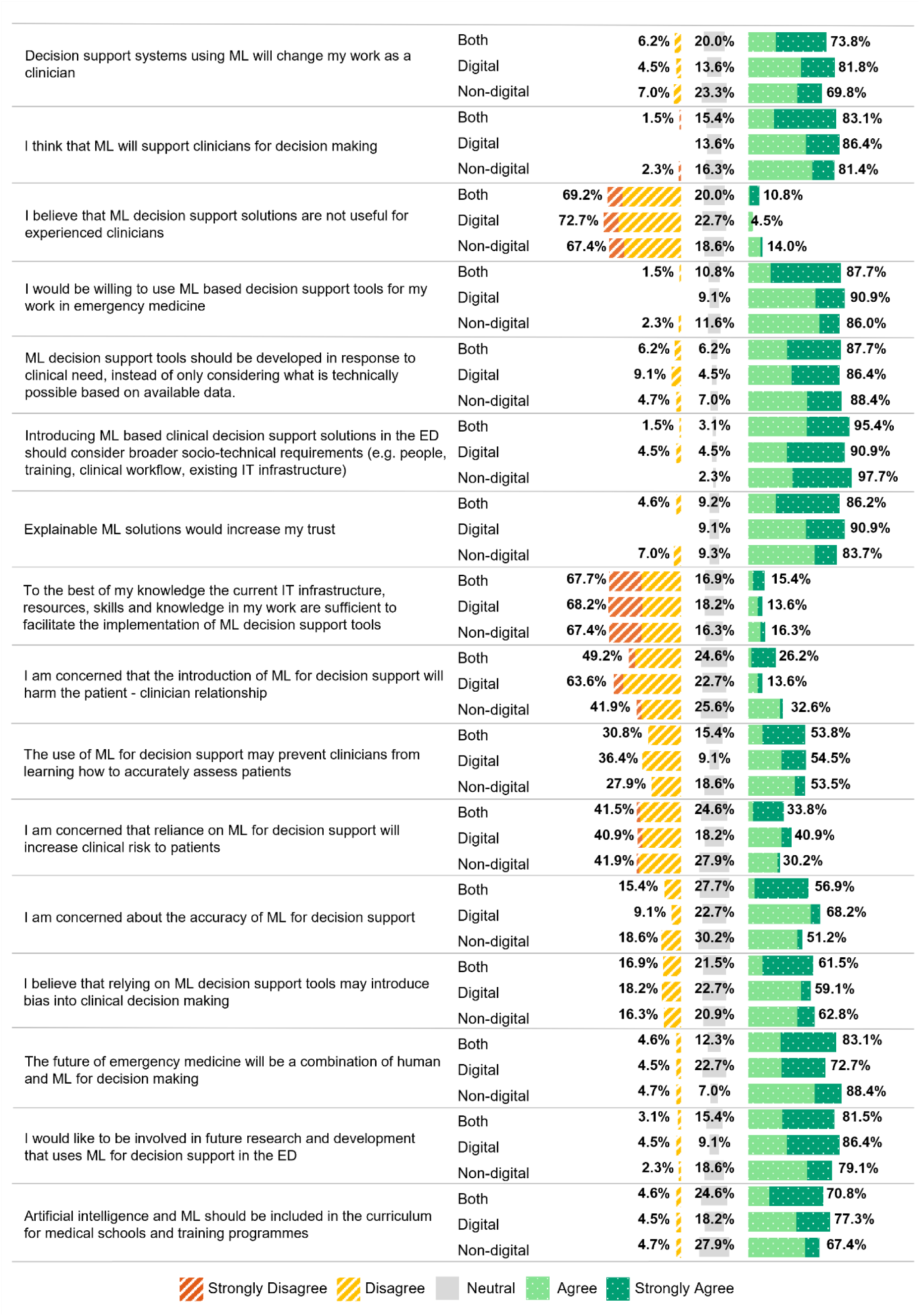
Perception, concerns, and the future of machine learning for decision support by digital and non-digital lead group. Abbreviations: ML, Machine Learning; ED, Emergency Department; IT, Information Technology

Fifty two respondents (80.0%) indicated that ML tools were not currently implemented, and were subsequently presented with statements exploring potential reasons. The majority (69.2%, 36/52) agreed that a lack of skilled resources is a significant issue. Concerns about insufficient or poor-quality electronic data were also common, with 55.8% (29/52) agreeing that this is a deterrent. Difficulty in deciding which processes would benefit most from ML solutions was a strong factor (digital leads: 43.8% (7/16), non-digital leads: 58.3% (21/36)). Digital leads were more convinced of the value of ML tools for emergency medicine (0%, 0/16 disagreed), whilst 30.6% (11/36) of non-digital leads were sceptical about their value. Lack of trust was higher among non-digital leads (11.1%, 4/36) compared to digital leads (0%, 0/16). ML models’ explanatory capabilities were considered underdeveloped (36.5%, 19/52), but most (65.4%, 34/52) disagreed that ML tools are too difficult to integrate into clinical workflows (Fig 3).

**Fig 3.**
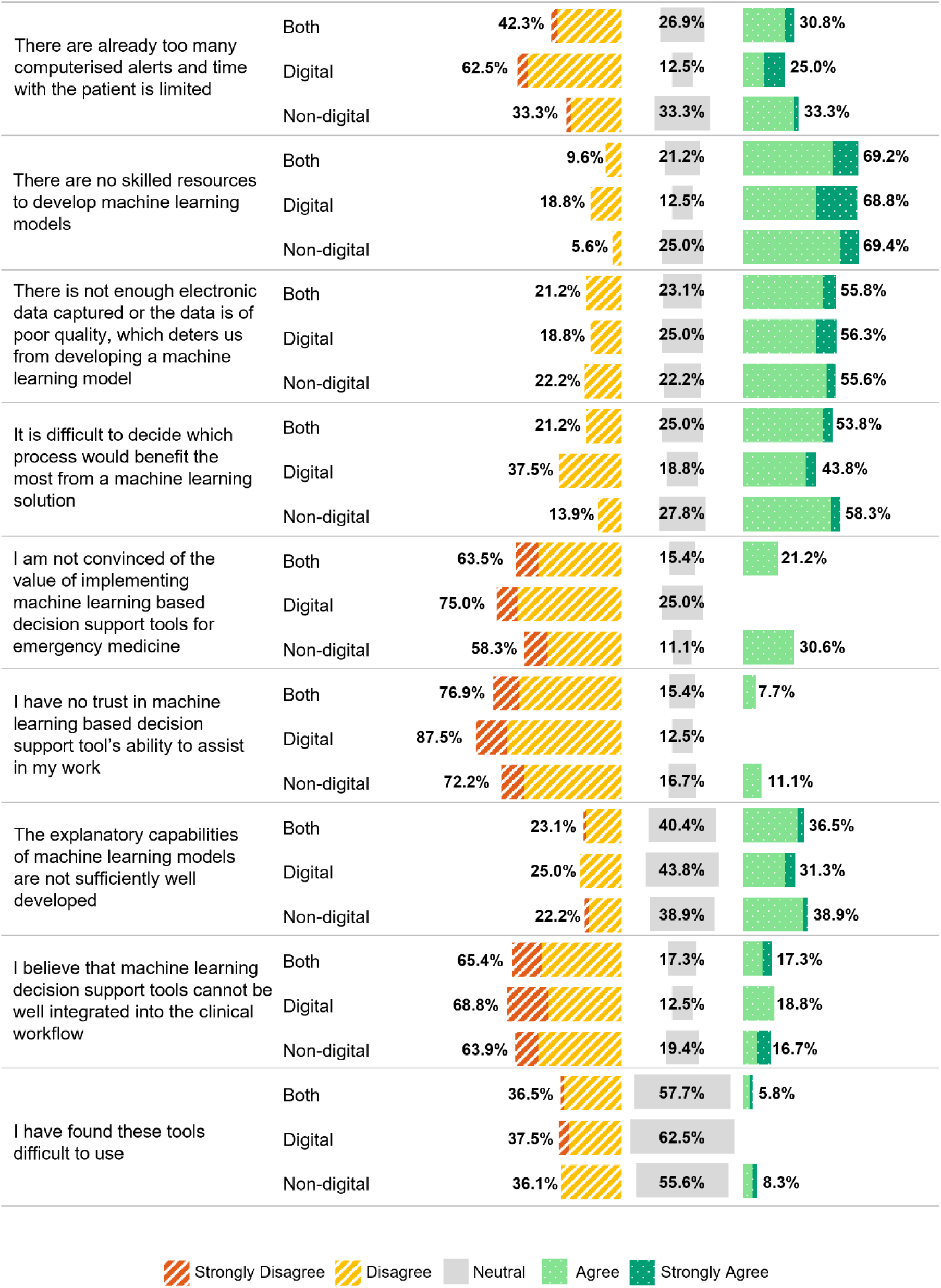
Opinion on why machine learning decision support tools are not used in the respondents clinical setting by digital and non-digital lead group.

The highest-rated applications of ML for decision support across both groups were antimicrobial stewardship (90.8%, 59/65), ML-assisted interpretation of electrocardiograms (ECG) (89.2%, 58/65), and analysis of radiology images (87.7%, 57/65). Lower-rated areas included diagnosis of mental health conditions (24.6%, 16/65) and patient experience and (dis)satisfaction prediction (35.4%, 23/65). (Fig 4).

**Fig 4.**
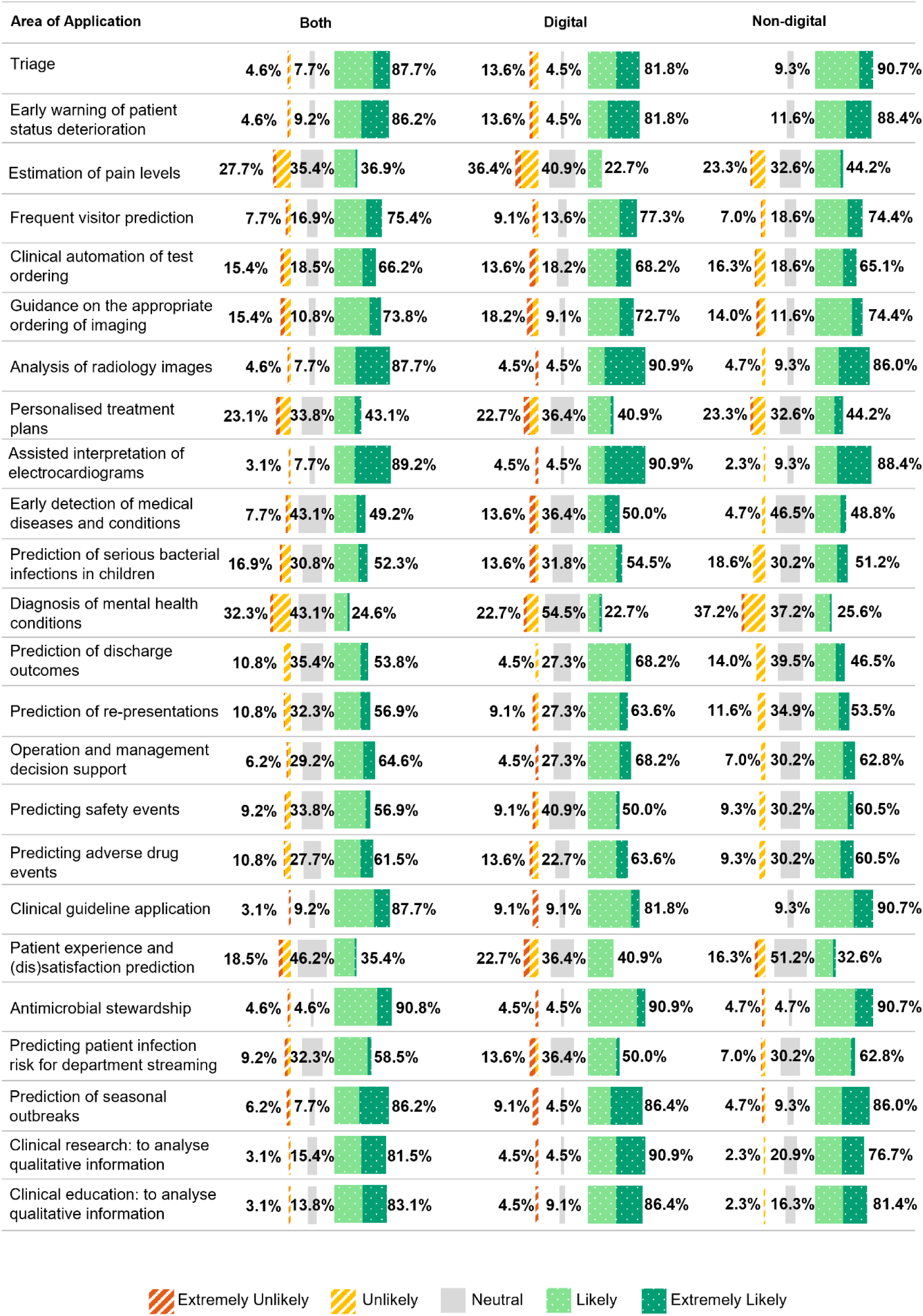
Areas of application of machine learning for decision support by digital and non-digital lead group.

Three respondents in the digital lead group (4.6%) confirmed that they had training in ML. One had accredited training and the other two were non-accredited or casual training. Most (89.2%, 58/65) expressed an interest in furthering their knowledge in ML.

There was a strong consensus among the respondents, particularly digital leads, on the value of sharing anonymised patient data for research (digital leads: 95.5% (21/22), non-digital leads: 90.7% (39/43)). Most (86.2%, 56/65) expressed interest in both accessing and contributing data to a cross-site data repository to facilitate research. Concerns about data protection were lower among digital leads, with 13.6% (3/22) expressing reluctance to share data, compared to 18.6% (8/43) of the non-digital leads (Fig 5). When asked about the extent of patient data the respondents would be willing to share to support a cross-site data repository for research purposes (once the appropriate approvals were in place), 44.6% (29/65) indicated they would be willing to share all or most of their data, 18/65 would contribute some, 17/65 were unsure and one would not contribute any.

**Fig 5.**
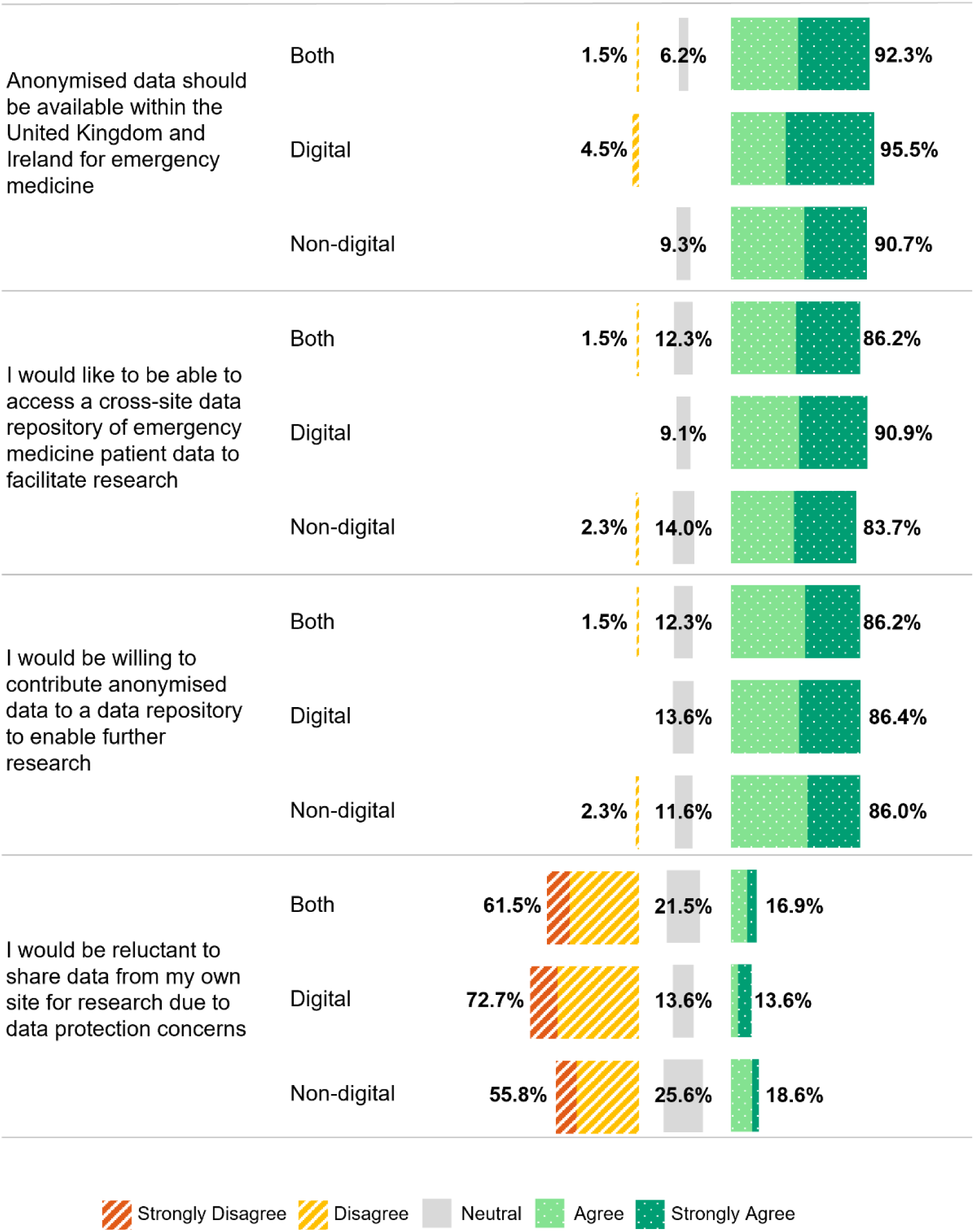
Assertions in relation to the sharing of fully anonymised patient data for research purposes by digital and non-digital lead group.

## Discussion

With the rapid growth in AI research within healthcare and the advent of advanced AI applications like ChatGPT [24], this survey comes at a pivotal time. To our knowledge, this is the first in children’s emergency care providing insights from frontline clinicians (specifically PERUKI site and digital leaders). A high response rate supports representation of a wide range of ED settings across the UK and Ireland. We have demonstrated both promise and challenges associated with integrating ML decision support tools into paediatric emergency care. While there is strong interest and willingness among clinicians to adopt these tools, gaps in AI literacy, infrastructure limitations, model accuracy, and concerns about bias present significant barriers. The educational video effectively reduced knowledge gaps, suggesting targeted training can improve clinician’s confidence in understanding key AI concepts, an essential step in adopting ML decision support tools. Digital leadership emerged as especially valuable in settings with various levels of digital maturity. ML tools were perceived to be most beneficial when based on clinical need. Furthermore, broad support for collaborative data sharing reflects an opportunity to build foundations for future research and innovation, provided concerns around data protection and security are addressed. These insights highlight the need for strategic investments in training, infrastructure, and leadership to ensure safe and equitable deployment of ML tools in clinical practice.

### Current knowledge, education and training

AI literacy involves understanding the fundamental functions of AI and how to utilise these applications effectively and responsibly [25]. The focus for most medical professionals is therefore about interacting with and using AI in healthcare rather than developing AI technologies or conducting AI research [26]. Understanding basic AI concepts is a key first step, as AI illiteracy can be a significant obstacle to adoption [27], especially for key decision makers. In this study digital leads had a stronger understanding of all AI concepts before watching the video, likely due to greater exposure to digital systems. The video narrowed the gap in AI knowledge between groups, with non-digital leads showing greater improvement in understanding advanced concepts like natural language processing and deep learning. This highlights the need to integrate targeted AI education into professional development, ensuring all clinical leaders, regardless of prior experience, are equipped to make informed decisions about integrating ML in healthcare for decision support.

Scott et al [28] advise that an important strategic enabler for adoption is enhancing clinician AI literacy, including AI/ML tool design and evaluation to assess suitability and appropriate use. Most respondents valued education and training, expressing interest in furthering their knowledge (89.2%), and supporting its inclusion in medical school curricula and training programmes (70.8%). Researchers believe that medical education needs to expand beyond the foundational biology, clinical skills and new diagnostic and therapeutic trends, with a key focus on preparing students to thrive in a healthcare system increasingly influenced by advancements in AI [29]. According to Ng et al [25] and also inspired by Bloom taxonomy, AI literacy should include fundamental competencies such as knowledge and understanding, usage and application, development and evaluation, along with social responsibility and ethical awareness. Most respondents (81.5%) would like to be involved in future research and development of ML based decision support tools. This co-design is essential for clinical utility, and education in AI/ML can enhance collaboration between clinicians and data scientists. However, the knowledge gap between clinicians, engineers, and scientists continues to grow as technology in healthcare advances, leaving physicians ill-prepared to work with AI tools [30] and impacting future collaboration. Medical schools should explore ways of introducing ML training modules to understand the promise and potential pitfalls [31].

### Opportunities and challenges

Most respondents (83.1%) believed these tools would enhance clinical decision making, with 87.7% willing to adopt them. However, concerns remain, the greatest relating to explainable AI (XAI), with 86.2% agreeing this would increase trust. XAI provides the ability for human experts to understand explanatory factors behind AI decision making, using established methods such as SHAP, LIME, and integrated gradients [32]. However, true understanding requires not just technical explanations, but also the ability of humans to comprehend them. Achieving this requires new human-AI interfaces enabling domain experts to ask questions and interactively explore explanations [33]. Another significant concern was bias being introduced into clinical decision making (61.5%). Bias in healthcare may arise from systemic inequalities embedded in healthcare data [34], but occurs in most stages of ML development: formulation of the research problem, data collection, data pre-processing, model development and validation and model implementation [35], leading to inequitable decision making, potentially impacting treatments and outcomes. A set of principles: findable, accessible, interoperable and reusable (FAIR) that were originally intended to be applied to data are now being designed for use with AI models [36] to address bias. Removing bias requires consideration of several areas, within model design, training data, and interactions with both clinicians and patients [37].

Accuracy of these tools was another concern. ML tools deployed in production must go through rigorous validation and have a proven level of accuracy that complies with clinical safety standards. Labkoff et al [38] through engagement with many diverse stakeholders produced guidelines for the development, validation and deployment of safe and trustworthy AI systems in healthcare. As patterns in data change over time, continuous monitoring and improvement of these models is important in the model lifecycle to detect performance degradation. ML Operations (MLOps) streamlines the ML lifecycle, integrating ML, software engineering, and data engineering, and should be considered essential for scalable, reliable, and efficient model deployment in healthcare [39]. Implementing MLOps strategies could mitigate concerns around clinical risk to patients with 33.8% of respondents agreeing this was a concern. With the European Union (EU) AI Act, high risk AI systems are required to set up post-market monitoring systems that will address any risks emerging from these systems to be addressed in a timely manner and as a legal requirement to report any breaches [40].

Many respondents (53.8%) believed using ML tools may prevent clinicians from learning how to accurately diagnose patients. It has been argued that incorporating ML decision support tools in medicine may lead to overreliance on automation, resulting in deskilling and curbing clinicians’ ability to learn, with reduced ability to make informed decisions when technology breaks down or fails [41]. However, this deskilling can be a result of clinicians’ own choices and external pressures of their work environment [42]. These tools are intended to support, not replace clinical judgement. The EU AI Act emphasises human oversight throughout the process for high risk AI systems and it is essential to ensure clinicians remain central in decision-making, understanding the limitations of the system, either disregarding or reversing outputs, and safely intervening and disabling the system when necessary [40,43].

Lastly, 67.7% believed current IT infrastructure, resources, skills, and knowledge were insufficient. Digital maturity levels across sites varied, with only half having fully electronic systems. Widespread implementation of AI requires significant IT system reconfiguration and supporting resources, which can be challenging in terms of securing investment amid competing healthcare priorities [7]. Interestingly, AI maturity models are now emerging to support the digital and AI maturity roadmap. These maturity models are useful tools in defining what degree of readiness an organisation is at in order to take advantage of AI [44].

### Potential applications

Among potential ML applications, antimicrobial stewardship, which focuses on appropriate antibiotic use was ranked one of the highest by both digital and non-digital leads at 90.8%. Antibiotics are often overprescribed in paediatric emergency medicine, partly due to clinician-caregiver interactions and diagnostic uncertainty, leading to risks such as antimicrobial resistance, adverse drug events and an increase in healthcare costs [45]. ML tools have the potential to reduce time spent reviewing data by anti-microbial team members, making predictions in seconds, freeing up time to focus more on diagnostic and therapeutic decisions instead [46]. In fact, the World Health Organization highlighted the importance of these programmes by publishing a practical toolkit in 2019 for low and middle income countries [47]. The paediatric emergency medicine priority setting partnership between PERUKI and the James Lind Alliance also listed how to safely reduce antibiotics use for children in their research priority list across the UK and Ireland [48]. The second highest ranked application was ML assisted interpretation of ECGs (89.2%), these tools could improve clinician interpretation of ECGs, reduce errors and expedite patient care in Eds [49]. The application ranked lowest by both groups at 24.6% was ML assisted diagnosis of mental health conditions. The number of paediatric patients presenting to the ED with mental health conditions is increasing globally, further straining already limited ED resources [50]. Shatt et al [51] carried out a scoping review of ML in mental health, with some promising applications that included detection and diagnosis of conditions such as depression, psychosis, schizophrenia and suicidal ideation, some of which included imaging data. However, diagnosis of these conditions may often take place in the community, outpatient or other specialised psychiatric settings rather than the ED.

### Data sharing

The accuracy and generalisability of AI models are strongly influenced by the availability of large, diverse and representative training data reflecting the target population [52]. Sharing fully anonymised patient data to create a repository would greatly enhance research and innovation for ML decision support tool development across the UK and Ireland. Maassen et al [17] identified that 82.5% of surveyed German hospital physicians agreed on the value of making data available for research, compared to 92.3% from our results. Willingness to contribute anonymised data and access it was exceeded in our study, suggesting that efforts to develop a research data sharing framework would be broadly supported. Data protection concerns were evident, which also surfaced when asked how much data they were willing to share, 27.7% would share some and 27.7% remained unsure or unwilling. This indicates a need for further reassurance and education on data security, in particular sophisticated techniques that can balance privacy protection with data utility [53], ensuring trust in data sharing. Federated learning, where ML models are trained on decentralised data, offers a promising alternative to centralised data sharing while addressing privacy concerns [38].

### Digital versus non-digital leads

Digital lead positions existed at 60% of the sites, indicating progress and investment in digital transformation. Digital leads demonstrated a higher initial understanding of key AI concepts, greater confidence that ML will impact their clinical work, a desire to participate in research and development, and a stronger belief that AI and ML should be included in medical school curricula. They also exhibited fewer trust issues, recognised the value of implementing these tools for decision support, showed greater willingness to share data, and had fewer data protection concerns. In contrast, non-digital leads showed greater education benefits from the educational video, were more convinced that the future would be a human-ML collaboration for decision making, however expressed more concern about harm to the patient-clinician relationship, clinical value, trust, explanatory capabilities and potential bias. Some of these differences likely reflect the digital leads familiarity with the technology and its implementation. This confirms the need for broader AI education to bridge knowledge gaps, build trust, and foster collaboration in adopting AI tools in clinical practice for all clinicians.

### Success factors for machine learning development

Success depends on selecting the right use cases with clear clinical value, Ramgopal et al [54] reports that AI clinical decision systems should answer clinically relevant questions. Engaging clinicians early in the research and development of AI tools ensures they are both practical and aligned with real-world clinical needs. Collaboration between clinicians, data scientists, and digital leaders is essential to the co-design process, as it fosters the development of more trustworthy and responsible XAI tools for high-stakes healthcare decisions [55], ultimately enhancing clinical adoption and improving patient outcomes. Increasing training and awareness is essential to bridge knowledge gaps, reduce resistance, and build trust in AI systems. Addressing common concerns such as explainability, bias, and patient safety is equally critical. Ensuring data quality and improving electronic health record infrastructure will enhance model performance. Finally, seamless clinical workflow integration is key to ensuring these tools support rather than disrupt clinical practice, enabling a more efficient human-ML partnership.

### Limitations

The survey was distributed through the PERUKI network, eliciting one response per site, reflecting the opinions of the leads as opposed to the wider ED workforce. Findings depended on the accurate self-reporting from a single respondent per site. Some respondents struggled with AI concepts that were explained in the educational video, evident from the low percentage of respondents who confirmed using knowledge (rule) based systems at 38.5%. Fewer digital leads compared to non-digital responded potentially impacting on responses around AI readiness and infrastructure challenges. Finally, the variability in digital maturity across the sites would have influenced responses, however this was also an important insight.

## Conclusion

This study highlights a strong clinician interest to integrate ML decision support tools into clinical workflows, recognising the benefits to decision making. Challenges remain, including infrastructure readiness, explainability, trust, concerns about bias, potential clinician deskilling and how humans and AI can work effectively together, highlighting the need for careful implementation strategies and ongoing human-AI collaboration. The digital leadership role is recognised as important in bridging the gap in knowledge, promoting AI adoption and guiding the implementation of digital and AI maturity within their site. Educational interventions can enhance AI literacy. Widespread support of collaborative data sharing presents important possibilities to create cross-site research frameworks to advance ML tool development. Maximising ML’s potential in decision support requires investment in training, infrastructure, and governance. Prioritising clinician engagement, responsible AI design, and seamless integration will ensure these tools enhance patient care. To further this research a survey should be carried out of the wider ED workforce.

## Supporting information

S1 File. Survey Questionnaire. Details of study participant information and the survey questions.

S2 File. Survey Checklist. Checklist for Reporting Results of Internet E-Surveys (CHERRIES).

## Data Availability

The data are not publicly available due to confidentiality and anonymisation commitments made to participants as outlined in the study's ethical approval. We are able to share aggregated results upon reasonable request.

## Acknowledgements

The authors would like to acknowledge Stewart McKenna, Children’s Health Ireland at Tallaght for carrying out survey usability and technical testing. We are also grateful to the research and innovation office in Children’s Health Ireland for providing access to develop the survey in REDCap and technical administrator support. Also acknowledged are the PERUKI site leads who distributed and for some sites also completed the survey: Meriel Tolhurst-Cleaver, Alder Hey Children’s Hospital NHS Foundation Trust, Liverpool; Daniel Murrell, Bedfordshire Hospitals NHS Foundation Trust - Luton and Dunstable University Hospital; Jonathan Adamson, Birmingham Children’s Hospital; Charlotte Munday, Bristol Royal Hospital for Children; Katherine Thompson, Chelsea and Westminster NHS Foundation Trust; Michael Barrett, Children’s Health Ireland at Crumlin; Sheena Durnin, Children’s Health Ireland at Tallaght; Patrick Fitzpatrick, Children’s Health Ireland at Temple Street; Emma Fauteux, Cork University Hospital; Hannah Walsh, Derriford Hospital, Plymouth; Darren Ranasinghe, Epsom General Hospital; Slyvester Gomes, Evelina London Children’s Hospital; Patrick Aldridge, Frimley Park Hospital; Mark Anderson, Great North Children’s Hospital, Newcastle Upon Tyne; Phil Peacock, Great Western Hospital, Swindon; Simon Richardson, Hull Royal Infirmary; David Hartin, Ipswich Hospital; Arshid Murad, James Cook University Hospital, Middlesbrough; Nicholas Richens, John Radcliffe Hospital, Oxford; Rachael Mitchell, King’s College Hospital London; Atif Latif, Kingston Hospital NHS Foundation Trust; Alice Downes, Leeds General Infirmary; Shane Fitzgerald, Leicester Royal Infirmary; Claire Kirby, Newham Hospital, London; Edward Snelson, Norfolk & Norwich University Hospitals; Neha Jain, North Middlesex Hospital; Pete Figg, Northern Devon Healthcare NHS Trust; Lee Tubman, Northumbria Healthcare NHS Foundation Trust; Paul Tanto, Northwick Park Hospital; Christopher Gough, Nottingham University Hospitals NHS Trust; Sharryn Gardner, Ormskirk & District General Hospital; Alan Charters, Queen Alexandra Hospital, Portsmouth; Gareth Patton, Royal Aberdeen Children’s Hospital; Michaela Lazner, Royal Alexandra Children’s Hospital, Brighton; Tom Waterfield, Royal Belfast Hospital for Sick Children; Manish Thakker, Royal Berkshire NHS Foundation Trust; Graham Johnson, Royal Derby Hospital; Hannah Stewart, Royal Devon and Exeter Hospital; Shye Wei Wong, Royal Free Hospital, London; Jen Browning, Royal Hospital for Children & Young People, Edinburgh; Steven Foster, Royal Hospital for Children, Glasgow; David Kung, Royal Manchester Children’s Hospital; Kirsty Challen, Royal Preston Hospital; Lorna Bagshaw, Royal Wolverhampton NHS Trust; Stephen Davies, Salisbury NHS Foundation Trust; Adrian Marsh, Shrewsbury & Telford NHS Trust; Esther Wilson, Somerset Foundation Trust Musgrove Hospital; Niall Mullen, South Tyneside & Sunderland NHS Foundation Trust; Alasdair Moffat, Southampton Children’s Hospital; Ellie Day, Southmead Hospital, North Bristol Trust; Heather Jarman, St George’s University Hospitals NHS Foundation Trust; Vanessa Merrick, St Mary’s Hospital, Imperial College Healthcare NHS Trust; Greg Cranston, The Grange Hospital, Newport; Emre Basatemur, The Royal London Hospital; Holly Brooker, Torbay and South Devon NHS Foundation Trust; Tulsi Patel, University College London Hospital; James Foley, University Hospital Galway; Marylyn Emeda, University Hospital Lewisham; George Simpson, University Hospital of North Tees; Michael Fox, University Hospital of Wales, Cardiff; Tadgh Moriarty, University Hospital Waterford; Katherine Priddis, Watford General Hospital; Rachel Shute, West Suffolk; Davin Amin, Wexham Park Hospital; Amutha Anpananthar, Whipps Cross Hospital, London; Alex Brown, Barnet Hospital; Catherine Williams, Bolton NHS Foundation Trust;

