## Supplementary material for "Machine Learning for Paediatric Related Decision Support in Emergency Care – A UK and Ireland Network Survey Study": S1 File. Survey Questionnaire. Details of study participant information and the survey questions.

### Machine Learning for Clinical Decision Support in Paediatric Emergency Medicine - Lead for Digital Systems

Please note that the institution that you currently work in should be reflected in the answers to this survey.

---

Study Participation Information

What is the purpose of this survey?

This survey presents an opportunity to gain valuable insight at site level into the current use, perceptions, concerns and the future need for machine learning clinical decision support system tools in the Emergency Department for paediatric patients across the United Kingdom and Ireland. Although the number of studies using machine learning for potential clinical decision support system tools is increasing each year, few surveys of clinicians on the use of machine learning in the emergency department exist, therefore this study comes at a pivotal time.

Please note that there are two surveys on this topic being distributed, this survey has been designed to be completed by the lead for digital systems (or nearest equivalent) in each Emergency Department site, the second should be completed by all other respondents that do not have this role. Only one version of the survey should be completed per person.

We aim to determine the use of clinical decision support system tools and machine learning in paediatric emergency medicine via an online survey, specifically:

Current information techology investment both in terms of infrastructure and clinician role/responsibility, recognising the value of data for decision making. Knowledge, attitudes and concerns around clinical decision support system tools. Suggested areas for development and future impacts. This survey is being performed as part of an academic qualification and will form one of the work packages for a PhD in data analytics and healthcare.

Why am I being asked to take part?

The adoption of machine learning decision support tools in healthcare will be largely driven by clinicians, yet little data exists regarding attitudes and knowledge of such technologies. It is also important to understand what areas of application are most important to clinicians, ensuring that effort is not spent on development of low priority tools. Your participation will contribute to this invaluable knowledge.

Do I have to take part?

No, your participation is completely voluntary and you will not be asked for your name or for any other identifying information.

If I take part, what will I be asked to do?

Once you have read this information leaflet and you are happy to proceed, you will then be asked five questions on your current knowledge of Artifical Intelligence key concepts. After which you will be asked to watch a short video that provides definitions of the core concepts and some examples of clinical decision support systems. You will then be asked to answer a short questionnaire, which should take approximately 8 minutes to complete.

Will the data be confidential?

Yes, this survey is completely anonymous. Your answers will be treated confidentially and the information you provide will be anonymised within any publications or reports. Your anonymised data will be held securely and can only be accessed by the investigators. All data will be deleted when the study and any subsequent publications or reports are completed, this usually takes about 5 years.

Any Questions?

If you have any questions or concerns on any aspect of this study, you can contact Fiona Leonard.

Please select 'Yes' to each statement if you understand and agree:

---

I have read and understand the Study Participant Information section.

◯ Yes

---

I understand that this survey is intended to be anonymous and should not identify any particular individual.

◯ Yes

---

I understand that should I provide my name or other identifiable information, my responses may be linked to me.

◯ Yes

---

I understand that if I include any identifiable information, these will be anonymised by the researchers.

◯ Yes

---

I understand that on completion of this survey I am providing consent for the use of this data.

◯ Yes

---

I understand that on completion of this survey, as everything is anonymised it will not be possible to have my specific responses removed from the pooled data.

◯ Yes

#### Participant Demographics

Site Location

- ◯ Addenbrooke's Hospital, Cambridge
- ◯ Alder Hey Children's Hospital NHS Foundation Trust, Liverpool
- ◯ Barking, Havering & Redbridge University Hospitals NHS Trust
- ◯ Barnet Hospital
- ◯ Bedfordshire Hospitals NHS Foundation Trust
- ◯ Birmingham Children's Hospital
- ◯ Bolton NHS Foundation Trust
- ◯ Bradford Royal Infirmary
- ◯ Bristol Royal Hospital for Children
- ◯ Chelsea and Westminster NHS Foundation Trust
- ◯ Children's Health Ireland at Crumlin
- ◯ Children's Health Ireland at Tallaght
- ◯ Children's Health Ireland at Temple Street
- ◯ Cork University Hospital
- ◯ Countess of Chester NHS Foundation Trust
- ◯ County Durham & Darlington NHS Foundation Trust
- ◯ Derriford Hospital, Plymouth
- ◯ Epsom General Hospital
- ◯ Evelina London Children's Hospital
- ◯ Frimley Park Hospital
- ◯ Gloucester Royal Hospital
- ◯ Great North Children's Hospital, Newcastle Upon Tyne
- ◯ Great Western Hospital, Swindon
- ◯ Ipswich Hospital
- ◯ Hull Royal Infirmary
- ◯ James Cook University Hospital, Middlesbrough
- ◯ John Radcliffe Hospital, Oxford
- ◯ King's College Hospital London
- ◯ Kingston Hospital NHS Foundation Trust
- ◯ Leeds General Infirmary
- ◯ Leicester Royal Infirmary
- ◯ Morriston Hospital, Swansea
- ◯ Newham Hospital, London
- ◯ Norfollk & Norwich University Hospitals
- ◯ North Middlesex Hospital
- ◯ Northern Devon Healthcare NHS Trust
- ◯ Northumbria Healthcare NHS Foundation Trust
- ◯ Northwick Park Hospital
- ◯ Nottingham University Hospitals NHS Trust
- ◯ Ormskirk & District General Hospital
- ◯ Peterborough City Hospital
- ◯ Queen Alexandra Hospital, Portsmouth
- ◯ Queen Elizabeth Hospital, Woolwich
- ◯ Royal Aberdeen Children's Hospital
- ◯ Royal Alexandra Children's Hospital, Brighton
- ◯ Royal Belfast Hospital for Sick Children
- ◯ Royal Berkshire NHS Foundation Trust
- ◯ Royal Derby Hospital
- ◯ Royal Devon and Exeter Hospital
- ◯ Royal Free Hospital, London
- ◯ Royal Hospital for Children, Glasgow
- ◯ Royal Hospital for Children & Young People, Edinburgh
- ◯ Royal Preston Hospital
- ◯ Royal United Hospital, Bath
- ◯ Royal Wolverhampton NHS Trust
- ◯ Salisbury NHS Foundation Trust
- ◯ Sheffield Children's NHS Foundation Trust
- ◯ Shrewsbury & Telford NHS Trust
- ◯ South Tyneside & Sunderland NHS Foundation Trust
- ◯ Southampton Children's Hospital
- ◯ St George's University Hospitals NHS Foundation Trust
- ◯ St Helen's & Knowsley NHS Trust
- ◯ St Mary's Hospital, Imperial College Healthcare NHS Trust

| | |
|---|---|
| | ◯ The Grange Hospital, Newport<br>◯ The Royal London Hospital<br>◯ Torbay and South Devon NHS Foundation Trust<br>◯ University College London Hospital<br>◯ University Hospital Crosshouse<br>◯ University Hospital Galway<br>◯ University Hospital Lewisham<br>◯ University Hospital of Wales, Cardiff<br>◯ University Hospital Waterford<br>◯ Walsall Manor Hospital<br>◯ Watford General Hospital<br>◯ West Suffolk<br>◯ Wexham Park Hospital<br>◯ West Middlesex Hospital<br>◯ Whipps Cross Hospital, London<br>◯ Whittington Health NHS Trust<br>◯ Other (United Kingdom)<br>◯ Other (Republic of Ireland) |
| Based on your selection of 'Other', please specify | ________________________ |
| Site Type | ◯ Paediatrics ◯ Paediatric and Adult<br>◯ Other |
| Based on your selection of 'Other' in the previous question, please describe further | ______________________________ |
| Respondent Type | ◯ Consultant<br>◯ Other |
| Based on your selection of 'Other' in the previous question, please describe further | ______________________________ |
| Clinician Type | ◯ Emergency Medicine<br>◯ Paediatrics<br>◯ Emergency Medicine with PEM subspecialty training<br>◯ Paediatrics with PEM subspecialty training<br>◯ Other |
| Based on your selection of 'Other' in the previous question, please describe further | ______________________________ |
| How many years have you worked in or with Emergency Medicine? | ◯ 0-2<br>◯ 3-5<br>◯ 6-10<br>◯ 10+ |

#### Are you confident that you understand the following Artificial Intelligence key concepts?

| | Strongly Agree | Agree | Neutral | Disagree | Strongly Disagree |
|---|---|---|---|---|---|
| I am confident that I understand the concept of artificial intelligence | ○ | ○ | ○ | ○ | ○ |
| I am confident that I understand the concept of machine learning within Artificial Intelligence | ○ | ○ | ○ | ○ | ○ |
| I am confident that I understand the concept of machine learning as it applies to 'image recognition' within Artificial Intelligence | ○ | ○ | ○ | ○ | ○ |
| I am confident that I understand the concept of natural language processing within Artificial Intelligence | ○ | ○ | ○ | ○ | ○ |
| I am confident that I understand the concept of deep learning within Artificial Intelligence | ○ | ○ | ○ | ○ | ○ |

**The following short video provides definitions of the core concepts in this survey, along with examples of clinical decision support systems. This will assist you in understanding some of the questions in the survey.**

**If required you can adjust the speed of the video to shorten the duration time by selecting the cog icon located at the bottom right of the video.**

Having watched the video do you still stand by the original answers you supplied for Artificial Intelligence and Machine Learning concept understanding? ○ Yes ○ No

Please provide your updated level of understanding of the following Artificial Intelligence key concepts.

| | Strongly Agree | Agree | Neutral | Disagree | Strongly Disagree |
| --- | --- | --- | --- | --- | --- |
| I am confident that I understand the concept of artificial intelligence | ○ | ○ | ○ | ○ | ○ |
| I am confident that I understand the concept of machine learning within Artificial Intelligence | ○ | ○ | ○ | ○ | ○ |
| I am confident that I understand the concept of machine learning as it applies to 'image recognition' within Artificial Intelligence | ○ | ○ | ○ | ○ | ○ |
| I am confident that I understand the concept of natural language processing within Artificial Intelligence | ○ | ○ | ○ | ○ | ○ |
| I am confident that I understand the concept of deep learning within Artificial Intelligence | ○ | ○ | ○ | ○ | ○ |

**In your opinion, please rate the following statements around perception, concerns and the future of ML for clinical decision support**

| | Strongly Agree | Agree | Neutral | Disagree | Strongly Disagree |
|---|---|---|---|---|---|
| Decision support systems using machine learning will change my work as a clinician | ◯ | ◯ | ◯ | ◯ | ◯ |
| I think that machine learning will support clinicians for decision making | ◯ | ◯ | ◯ | ◯ | ◯ |
| I believe that machine learning decision support solutions are not useful for experienced clinicians | ◯ | ◯ | ◯ | ◯ | ◯ |
| I would be willing to use machine learning based decision support tools for my work in emergency medicine | ◯ | ◯ | ◯ | ◯ | ◯ |
| Machine learning decision support tools should be developed in response to clinical need, instead of only considering what is technically possible based on available data. | ◯ | ◯ | ◯ | ◯ | ◯ |
| The introduction of machine learning based clinical decision support solutions in the ED should consider broader socio-technical requirements (e.g. people, training, clinical workflow, existing information technology infrastructure) | ◯ | ◯ | ◯ | ◯ | ◯ |
| Explainable machine learning solutions would increase my trust | ◯ | ◯ | ◯ | ◯ | ◯ |
| To the best of my knowledge the current information technology infrastructure, resources, skills and knowledge in my work are sufficient to facilitate the implementation of machine learning decision support tools | ◯ | ◯ | ◯ | ◯ | ◯ |
| I am concerned that the introduction of machine learning for decision support will harm the patient - clinician relationship | ◯ | ◯ | ◯ | ◯ | ◯ |

| | | | | | |
|---|---|---|---|---|---|
| The use of machine learning for decision support may prevent clinicians from learning how to accurately assess patients | ○ | ○ | ○ | ○ | ○ |
| I am concerned that reliance on machine learning for decision support will increase clinical risk to patients | ○ | ○ | ○ | ○ | ○ |
| I am concerned about the accuracy of machine learning for decision support | ○ | ○ | ○ | ○ | ○ |
| I believe that relying on machine learning decision support tools may introduce bias into clinical decision making | ○ | ○ | ○ | ○ | ○ |
| The future of emergency medicine will be a combination of human and machine learning for decision making | ○ | ○ | ○ | ○ | ○ |
| I would like to be involved in future research and development that uses machine learning for decision support in the Emergency Department | ○ | ○ | ○ | ○ | ○ |
| Artificial intelligence and machine learning should be included in the curriculum for medical schools and training programmes | ○ | ○ | ○ | ○ | ○ |

#### Current Information Technology set up, along with current experience with clinical decision support system tools

Are you the lead for digital systems in your department?

◯ Yes ◯ No

Does this position exist in your department?

◯ Yes ◯ No

Please explain further

__________________________________

Does your department utilise data from healthcare records to drive improvements?

◯ Yes ◯ No

In my current work, emergency department patient records are recorded as one of the following

◯ Paper based
◯ Paper and electronic system
◯ Fully electronic
◯ Other
◯ Unsure

Based on your selection of 'Other' in the previous question, please describe further

__________________________________

What kind of information system is emergency department patient data currently recorded on?

Abbreviations:

- Emergency Department Information System (EDIS)
- Electronic Healthcare Record (EHR)

☐ Standalone EDIS (independent system, specific to ED that does not talk to other systems)
☐ Integrated EDIS (ingests data from other systems, where data is captured independently on for example, the patient administration system or laboratory information management system)
☐ ED module within a hospital wide EHR System
☐ Other

Based on your selection of 'Other' in the previous question, please describe further

__________________________________

What other systems are integrated with the Emergency Department Information System?

☐ Patient Administration
☐ Outpatient
☐ Inpatient
☐ Pharmacy
☐ Laboratory
☐ Theatre
☐ Inventory
☐ Radiology
☐ Other clinical systems
☐ None of the above

Based on your selection of 'Other clinical systems' in the previous question, please describe further

__________________________________

I use knowledge based electronic decision support tools in my work

◯ Yes ◯ No ◯ Unsure

| | |
|---|---|
| Which knowledge based decision support tools have you used? | ☐ Triage<br>☐ Early Warning Scores<br>☐ Realtime Emergency Department dashboard<br>☐ Diagnostic support<br>☐ Image or test ordering systems (provides advice based on knowledge database)<br>☐ Alert mechanisms warning of potential drug-drug interactions<br>☐ Clinical guideline support for specific medical conditions<br>☐ Pain assessment or management<br>☐ Other<br>☐ None of the above |
| Based on your selection of 'Other or none of the above' in the previous question, please describe further | __________________________________ |
| Have you contributed to a project that uses machine learning in emergency medicine? | ○ Yes ○ No |
| Have you ever used machine learning (non-knowledge based) decision support tools in your clinical work? | ○ Yes ○ No |
| Which decision support purposes have you used these clinical support tools for? | __________________________________ |
| How often would you use these tools? | ○ Monthly ○ Weekly<br>○ Daily ○ Other |
| Based on your selection of 'Other or none of the above' in the previous question, please describe further | __________________________________ |
| There are no machine learning models deployed in my work for decision support | ○ True ○ False |

**In your opinion, machine learning decision support tools are not used in my clinical setting because:**

| | Strongly Agree | Agree | Neutral | Disagree | Strongly Disagree |
|---|---|---|---|---|---|
| There are already too many computerised alerts and time with the patient is limited | ◯ | ◯ | ◯ | ◯ | ◯ |
| There are no skilled resources to develop machine learning models | ◯ | ◯ | ◯ | ◯ | ◯ |
| There is not enough electronic data captured or the data is of poor quality, which deters us from developing a machine learning model | ◯ | ◯ | ◯ | ◯ | ◯ |
| It is difficult to decide which process would benefit the most from a machine learning solution | ◯ | ◯ | ◯ | ◯ | ◯ |
| I am not convinced of the value of implementing machine learning based decision support tools for emergency medicine | ◯ | ◯ | ◯ | ◯ | ◯ |
| I have no trust in machine learning based decision support tools ability to assist in my work | ◯ | ◯ | ◯ | ◯ | ◯ |
| The explanatory capabilities of machine learning models are not sufficiently well developed | ◯ | ◯ | ◯ | ◯ | ◯ |
| I believe that machine learning decision support tools cannot be well integrated into the clinical workflow | ◯ | ◯ | ◯ | ◯ | ◯ |
| I have found these tools difficult to use | ◯ | ◯ | ◯ | ◯ | ◯ |

In your opinion, are there any other reasons for not using machine learning decision support tools in your clinical setting? ________________________________

#### Training in Machine Learning

| | |
|---|---|
| Have you received any training in machine learning? | ◯ Yes ◯ No |
| What kind of training? | ☐ Accredited ☐ Non-accredited<br>☐ Casual |
| Do you have an interest in furthering your knowledge in machine learning? | ◯ Yes ◯ No |
| In your opinion, what do you think should be done to prepare clinicians for the introduction of machine learning for decision support in emergency medicine? | __________________________________ |

**Areas of application for Machine Learning in Emergency Medicine**

**Please rate, in your opinion, whether using Machine Learning could significantly enhance decision support for Emergency Medicine in the following areas of application.**

| | Extremely unlikely | Unlikely | Neutral | Likely | Extremely likely |
|---|---|---|---|---|---|
| Triage | ○ | ○ | ○ | ○ | ○ |
| Early warning of patient status deterioration | ○ | ○ | ○ | ○ | ○ |
| Estimation of pain levels | ○ | ○ | ○ | ○ | ○ |
| Frequent visitor prediction | ○ | ○ | ○ | ○ | ○ |
| Clinical automation of test ordering | ○ | ○ | ○ | ○ | ○ |
| Guidance on the appropriate ordering of imaging | ○ | ○ | ○ | ○ | ○ |
| Analysis of radiology images | ○ | ○ | ○ | ○ | ○ |
| Personalised treatment plans | ○ | ○ | ○ | ○ | ○ |
| Machine learning assisted interpretation of electrocardiograms | ○ | ○ | ○ | ○ | ○ |
| Early detection of medical diseases and conditions | ○ | ○ | ○ | ○ | ○ |
| Prediction model for serious bacterial infections in children | ○ | ○ | ○ | ○ | ○ |
| Diagnosis of mental health conditions | ○ | ○ | ○ | ○ | ○ |
| Prediction of discharge outcomes | ○ | ○ | ○ | ○ | ○ |
| Prediction of re-presentations | ○ | ○ | ○ | ○ | ○ |
| Operation and management decision support | ○ | ○ | ○ | ○ | ○ |
| Predicting safety events | ○ | ○ | ○ | ○ | ○ |
| Predicting adverse drug events | ○ | ○ | ○ | ○ | ○ |
| Clinical guideline application | ○ | ○ | ○ | ○ | ○ |
| Patient experience and (dis)satisfaction prediction | ○ | ○ | ○ | ○ | ○ |
| Antimicrobial stewardship | ○ | ○ | ○ | ○ | ○ |
| Predicting patient infection risk for department streaming | ○ | ○ | ○ | ○ | ○ |
| Prediction of seasonal outbreaks | ○ | ○ | ○ | ○ | ○ |
| Clinical research: to analyse qualitative information | ○ | ○ | ○ | ○ | ○ |

| | | | | | |
|---|---|---|---|---|---|
| Clinical education: to analyse qualitative information | ○ | ○ | ○ | ○ | ○ |

In your opinion, are there any other areas in emergency medicine that you think machine learning could assist with decision support? __________________________________

**Data Repository**

**A data repository containing anonymised patient data would allow the secondary use of medical information for research purposes, which would include the development of machine learning models.**

**Please rate the following assertions, in your opinion, in relation to the sharing of fully anonymised patient data for research purposes**

| | Strongly Agree | Agree | Neutral | Disagree | Strongly Disagree |
|---|---|---|---|---|---|
| Anonymised data should be available within the United Kingdom and Ireland for emergency medicine | ○ | ○ | ○ | ○ | ○ |
| I would like to be able to access a cross-site data repository of emergency medicine patient data to facilitate research | ○ | ○ | ○ | ○ | ○ |
| I would be willing to contribute anonymised data to a data repository to enable further research | ○ | ○ | ○ | ○ | ○ |
| I would be reluctant to share data from my own site/research due to data protection concerns | ○ | ○ | ○ | ○ | ○ |

Subject to the correct approvals, how much of your emergency medicine data is accessible for research?

- ○ None
- ○ Some
- ○ Most
- ○ All
- ○ Unsure
