## Supplementary material for "Machine Learning for Paediatric Related Decision Support in Emergency Care – A UK and Ireland Network Survey Study": S2 File. Survey Checklist. Checklist for Reporting Results of Internet E-Surveys (CHERRIES).

Eysenbach G. Improving the quality of Web surveys: the Checklist for Reporting Results of Internet E-Surveys (CHERRIES). J Med Internet Res 2004;6:e34

| **Checklist for Reporting Results of Internet E-Surveys (CHERRIES)** | | |
|---|---|---|
| ***Item Category*** | ***Checklist Item*** | ***Response*** |
| **Design** | Describe survey design | The target population was Emergency Departments that provide care to children and young people. The survey was distributed via the PERUKI network (a research network that includes England, Ireland, Northern Ireland, Scotland and Wales) made up of clinicians and academics. One response per site was sought, either the digital lead or if unavailable, the PERUKI site lead. |
| **IRB (Institutional Review Board) approval and informed consent process** | IRB approval | Ethical approval was received from both Children's Health Ireland (REC-281-23) and Technological University Dublin (LEWS-023-83). |
| | Informed consent | Participant information was included at the beginning of the survey. Participants were informed of the survey length (eight minutes, excluding the video at approximately three and a half minutes). Participants were informed that the survey was voluntary, completely anonymous, data would be stored securely, accessible only to the study investigators, and retained for approximately five years whereby it would be deleted. The participants were advised of the purpose of the study and who the investigators were. |
| | Data protection | No personal information was collected. |
| **Development and pre-testing** | Development and testing | The survey questions were informed by previous research and further input, discussion and consensus from the study investigators.<br>The survey was tested (usability and technical functionality) by the study investigators and a clinician who provided feedback as an independent reviewer, with adjustments and several revisions made based on feedback received. |
| **Recruitment process and description of the sample having access to the questionnaire** | Open survey versus closed survey | This was a closed survey, co-ordinated/distributed to individuals by the PERUKI site lead. |
| | Contact mode | Contact was through the PERUKI network, whereby the PERUKI site leads were emailed. |
| | Advertising the survey | The survey was not advertised. Targeted individual recruitment via the PERUKI site leads was facilitated through the PERUKI network. |

**Checklist for Reporting Results of Internet E-Surveys (CHERRIES)**

| ***Item Category*** | ***Checklist Item*** | ***Response*** |
|---|---|---|
| **Survey administration** | Web/E-mail | A link to the online survey developed in REDCap was sent via email. |
| | Context | REDCap is a secure online database for electronic data capture, with functions that include survey development and delivery (www.project-redcap.org). |
| | Mandatory/voluntary | This was a voluntary survey. |
| | Incentives | There were no incentives offered for participation. |
| | Time/Date | The duration of the survey was between 14th June 2024 and 12th July 2024. |
| | Randomization of items or questionnaires | For sections with many questions, random order was utilised. This included all questions utilising a matrix in REDCap. |
| | Adaptive questioning | Adaptive questioning was used throughout the survey to minimize questions asked to be only those relevant based on opening questions. |
| | Number of Items | Dependent on adaptive questioning, the maximum was 104 fields. |
| | Number of screens (pages) | The maximum number of pages (sections) dependent on adaptive questioning was 10. |
| | Completeness check | Questions requiring a response were tagged as mandatory, and respondents were asked to review and provide responses for any such fields prior to submission. |
| | Review step | Yes, respondents were able to review and change their answers prior to final submission by selecting the navigation buttons. |
| **Response rates** | Unique site visitor | The audit functionality of REDCap allows logging of visits and responses. This was combined with collection of site names to ensure a maximum of one visit (and response) per site was possible |
| | View rate (Ratio of unique survey visitors/unique site visitors) | Not applicable – respondents were invited via the PERUKI network. |
| | Participation rate (Ratio of unique visitors who agreed to | 67 responses from 75 invited (89.3% participation rate) |

**Checklist for Reporting Results of Internet E-Surveys (CHERRIES)**

| *Item Category* | *Checklist Item* | *Response* |
|---|---|---|
| | participate/unique first survey page visitors) | |
| | Completion rate (Ratio of users who finished the survey/users who agreed to participate) | 65 completed surveys out of a total of 67 responses (97% completion rate) |
| **Preventing multiple entries from the same individual** | Cookies used | Cookies were not used. Other methods, including naming of site and REDCap logging tools, were utilised. |
| | IP check | IP address was not used. Other methods, including naming of site and REDCap logging tools, were utilised. |
| | Log file analysis | Log file analysis was not used. Other methods, including naming of site and REDCap logging tools, were utilised. |
| | Registration | A login was not used. The invitation to the survey was via email that contained a web link. Duplicates were found based on the site name, however all duplicate site entries were incomplete (at most 3 pages of the survey were filled in), therefore they were excluded. |
| **Analysis** | Handling of incomplete questionnaires | Only completed responses were analysed and included. |
| | Questionnaires submitted with an atypical timestamp | No respondents were removed from the survey based on the time it took to complete. |
| | Statistical correction | Weighting of items or propensity scores were not used in the analysis of results. |
